# Relationship between Transposable Elements and behavioral traits: insights from six genetic isolates from North-Eastern Italy

**DOI:** 10.1101/2025.05.07.25327148

**Authors:** Giorgia Modenini, Giacomo Mercuri, Paolo Abondio, Giuseppe Giovanni Nardone, Aurora Santin, Paola Tesolin, Beatrice Spedicati, Alessandro Pecori, Giulia Pianigiani, Maria Pina Concas, Giorgia Girotto, Paolo Gasparini, Alessio Boattini, Massimo Mezzavilla

## Abstract

Half of the human genome is derived from Transposable Elements (TEs), among which Alu, LINE-1 and SVA are particularly represented. Germline transposition of TEs generates polymorphisms between individuals and may be used to study association with phenotypes and inter-individual differences. Italy presents an increased number of isolated villages compared to other European groups, and these isolates provide a desirable study subject to understand the genetic variability of the Italian peninsula. Therefore, we focused on the association between polymorphic TEs, behavioral traits (tobacco use and alcohol consumption), and Body Mass Index (BMI) variations, which could lead to an increased risk of developing addiction-related or metabolic diseases. We identified 12,709 polymorphic TEs in 589 individuals from six isolates: classical population genetics analyses showed that while closely related to other European populations, the isolates tend to cluster amongst themselves and are dominated by drift-induced ancestral components. Several TEs were also significantly associated with behavioral traits (tobacco use or alcohol consumption) or with BMI variations and some of them have a functional role. These results suggest that polymorphic TEs may significantly impact inter-individual and inter-population phenotypic differentiation, while also functioning as variability markers and potentially having a role in susceptibility to medical conditions.

## Introduction

The Italian peninsula, due to its complex population structure, could play an important role in the understanding of the genetic diversity of current populations, being the natural crossroad for human migrations across the Mediterranean since prehistoric periods. These migration patterns left a tangible mark on present-day Italians, revealing a heterogeneous network of genomic landscapes across the peninsula, with North Italian groups being more closely related to Western/Eastern European populations and a progressively increasing genetic connection with Northern African and Middle Eastern populations as we move southwards [1]. On top of this clinal variation across the peninsula, the natural variety of environments [2] provoked a series of local adaptive events that determined, among other factors, a differential disease susceptibility of Italian subpopulations [1]. A refined understanding of these local events would improve our knowledge of human diversity as a whole, and on a more practical level allow us to provide more *ad hoc* medical care and measures to particularly susceptible subpopulations.

The underlying genetic variability of Italy remains under-sampled and underrepresented, with available human genome reference datasets such as the 1KGP, HGDP, and SGDP only sampling three populations for the whole peninsula: Tuscans (TSI), individuals from Bergamo and Sardinians, a notion that only worsens when considering that Italy presents an increased number of isolated villages and subpopulations when compared to other European groups [3,4], most of which remain uncharacterized. These still isolated groups provide a desirable study subject to understand the Italian genetic variability: population isolates are characterized by small effective population sizes (Ne), which result in a decreased variability and stronger genetic drift effects, potentially increasing the frequency of variants that are rare or absent elsewhere and aiding at the discovery of novel rare variant signals underpinning complex traits such as medical risks and susceptibilities [5]. Population isolates tend not only to be genetically homogenous but are also characterized by an elevated diversity when compared to neighboring populations and their source population [3], because of geographical and/or cultural barriers that are necessary for the formation of the isolate in the first place. For these reasons, the isolates provided useful tools for genome-wide association studies [6].

However, most of the available research on these populations is based almost exclusively on SNP data, while little work was done using other types of genetic markers. For instance, information about the variability of Transposable Elements (TE), despite them being a primary component of the human genome, has become accessible only in recent years, thanks to the availability of whole-genome sequencing data and in particular to the development of new tools for their detection and genotyping [7–9]. When TEs transpose in the germline, they can create novel inheritable insertions, thereby generating human-specific polymorphisms [7]. One of the most useful features of polymorphic TEs is that the ancestral state of these markers is known to be the absence of the insertion [10,11]. Interestingly, such markers have never been used to study the genetic underpinnings of human isolated communities; therefore, this study is the first of a kind.

In the last decades, we have come to know much more about the impact of these elements on the genome and gene networks, and it has been shown that TE insertions can generate diversity in a variety of ways. For example, transposable elements have been linked to providing polyadenylation signals inducing the termination of transcripts [12], modifying splicing patterns, and providing new splicing sites [13], epigenetically affecting nearby genes [14,15], acting as novel promoters, enhancers, and transcription factor building sites [16,17], and often carrying their enhancers and promoters [18]. With their innate ability to act as disruptors and deregulators of gene expression, TE insertions have been associated with a variety of human diseases: for example, several cancer types [19,20], hemophilia A and B [21,22], some inheritable genetic diseases such as Dent’s disease or Duchenne muscular dystrophy [23], metabolic diseases [24], substance abuse, and central nervous systems diseases [25].

In particular, much interest has been given in recent years to the impact of transposable elements on the central nervous system [25–27]. Genome-wide approaches allowed researchers to study the role of transposable elements in stress-related learning mechanisms in rats [28], which have been used as a model for PTSD in humans [29]. Likewise, transposable elements have also been associated with alcoholism in humans using the same genome-wide approach [25].

In this study, we aim to reconstruct the TE variability of six isolates from Friuli-Venezia Giulia (North-Eastern Italy) thanks to the availability of whole sequencing data from 589 individuals [3]. Firstly, after determining the position and the genotypes of polymorphic TEs in these populations, we use them to evaluate the isolates’ structure, in the context of European and worldwide reference populations. Then, leveraging on the advantages offered by genetic and geographic isolates, we focused on exploring the potential association between non-reference polymorphic TEs, Body Mass Index (BMI) variations and behavioral traits of health and social relevance such as tobacco use and alcohol consumption. In fact, these traits could lead to an increased risk of developing addiction-related or metabolic diseases, such as tobacco use disorder, alcoholism, and obesity.

## Results

### TE variation distribution

After the analysis of polymorphic non-reference TEs with MELT v.2.2.2 [8], a total of 9,525 Alus, 2,283 LINE1s, and 901 SVAs were retrieved.

Then, allele frequencies were scanned for significant differences among the isolates: this way, a total of 3,987 TEs (31.37%) were identified as “differentiated”, of which 3,195 Alus (33.54%), 636 LINE1s (27.86%), and 156 SVAs (17.31%). When considering all comparison European populations, the corresponding rates of “differentiated” TEs are 53.45% (Alus), 58.63% (LINE1s) and 51.24% (SVAs). Of these insertions, we also considered their location (Table 1).

**Table 1.**
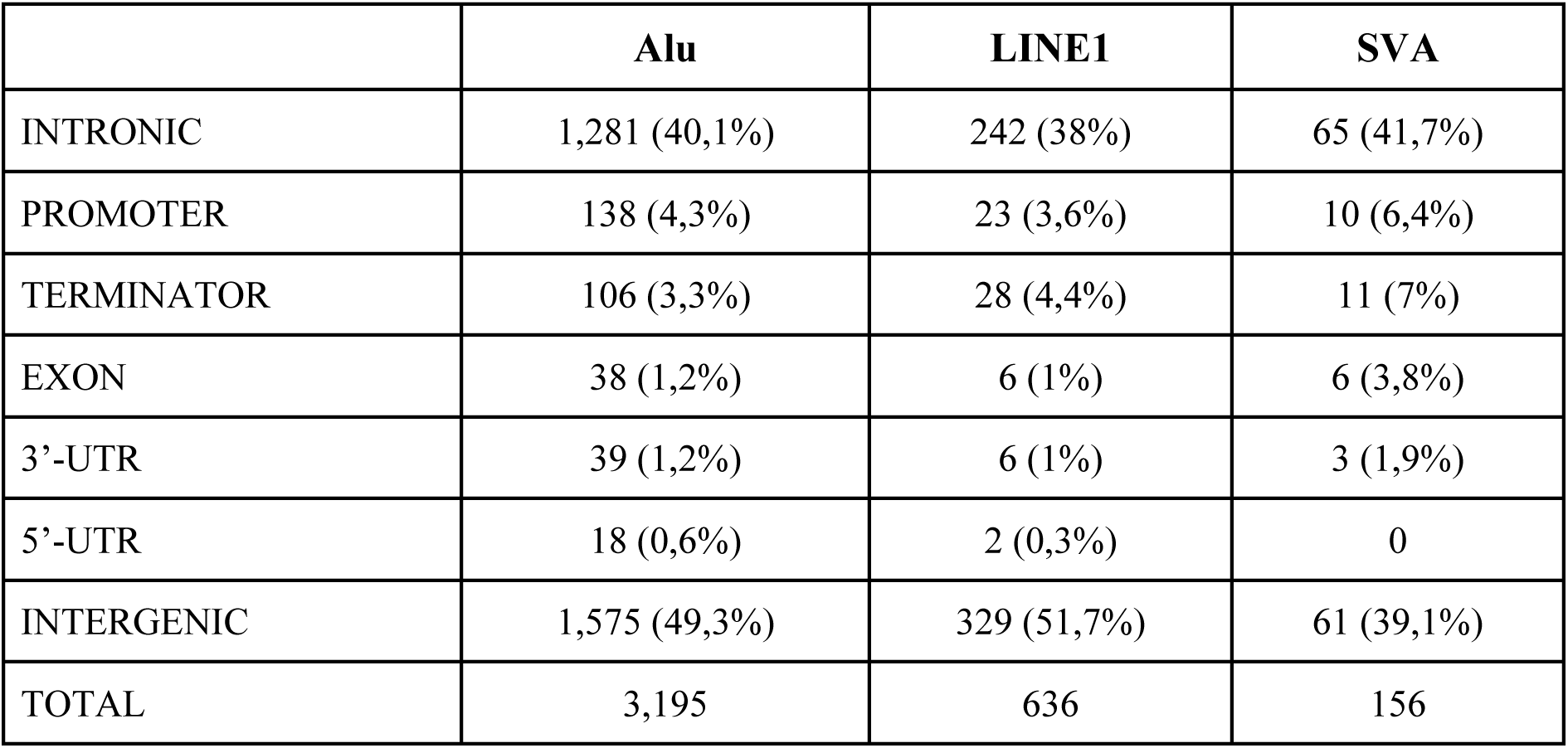
Significantly different polymorphic TEs between the six villages, divided by insertion location relative to gene region (with percentages) and TE superclass.

As expected, most polymorphic TE insertions are located in intronic and intergenic regions and only a negligible fraction are located in exonic regions (Supplementary Table S1). However, it is interesting to note that SVAs, which can be up to 3kb long [44], are overall less frequent in intergenic sequences while they appear more often located in “functional” regions (regulators or exons) when compared to Alus and LINE1s. This finding corroborates the notion that SVA insertions have the innate potential to regulate gene expression through their location insertion and their sequence characteristics [45,46].

### TE as markers for population structure

Both TE-based PCA and Admixture showed that, while closely related to other European populations, our isolates tend to cluster amongst themselves and are dominated by drift-induced ancestry components (Supplementary Figure S1). In particular, the first PC discriminates between African and non-African populations, while the second PC highlights a West-to-East geographical pattern including individuals from Friuli-Venezia Giulia, Europeans, Americans, South Asians, and East Asians.

The PCA between European and FVG populations divides the two groups along the first PC, while the second component highlights the variability between the isolates, separating Resia and some individuals from Clauzetto and Sauris from the rest (Figure 1A). As expected considering their geographical proximity and historical relatedness, Tuscans (TSI) and Central Europeans (CEU) are the closest groups to the FVG isolates. This PCA is similar to the one resulting in Esko et al. [3] based on SNPs. Looking at the second and third PCs, it is interesting to note that PC2 separates Resia from Clauzetto, while the third component highlights the differentiation between Sauris and Illegio. Instead, Erto, San Martino and most individuals from Clauzetto cluster together with the other European populations (Figure 1B), hence suggesting a lower degree of isolation for these groups. Finally, looking at the Admixture graph (Figure 1C), the “tidiest” model is for K = 7, as K = 9 presents excessive noise, especially in the African outgroup. However, at the best fitting K = 9 (CV error = 0.31088), Illegio, Resia, San Martino and Sauris are all dominated by their own ancestry components, which are present only marginally in Clauzetto, Erto, and the other European populations.

**Figure 1.**
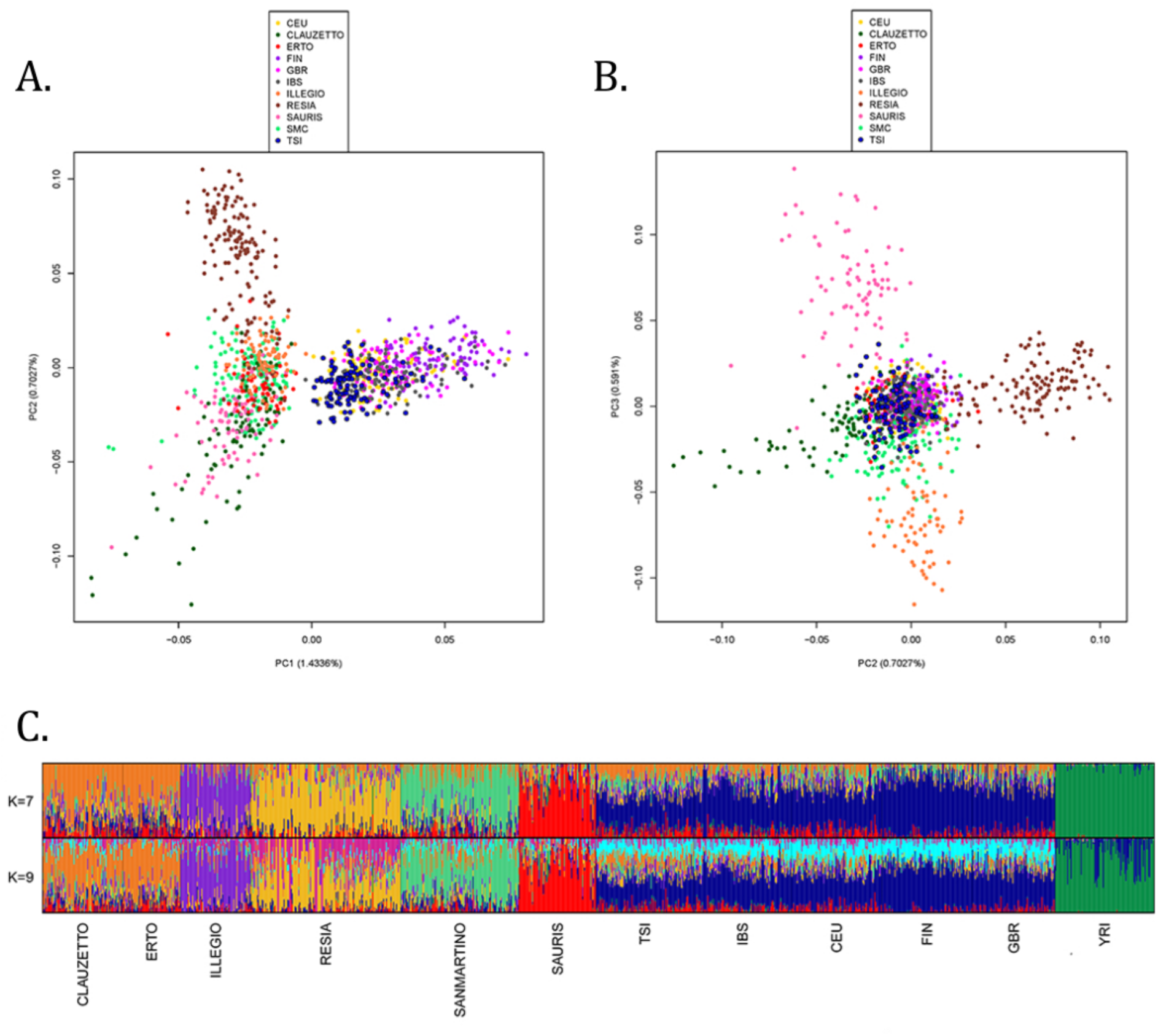
A and B) PCA plots of European populations from 1KGP and FVG isolates, first against second component (A) and second against third component (B). C) Admixture barplots for K = 7 and K = 9.

### Association between TEs and selected phenotypes

Several polymorphic TEs were identified by the association tests with GEMMA [35,36] as possibly associated with the conditions detailed in Materials and Methods, and some of them also act as eQTLs/sQTLs:

1. Variations in Body Mass Index: three insertions were deemed significant, namely twoAlus and one SVA (Figure 2, middle plot). Notably, the SVA on chr17:49150166 is located in the gene SPAG9 (Sperm Associated Antigen 9) (Table 2). The other two significant results are two intergenic Alus located on chr1:241980544 and chr14:65796449. Interestingly, both act as eQTLs and sQTLs in several tissues, such as pituitary, blood, heart, testis, and ovary.
2. Alcohol consumption: six Alus were found to be significant (Figure 2, inner Manhattan plot), with only one in a genic region, the Alu on chr12:14020945 in the gene *GRIN2B* (Glutamate Ionotropic Receptor NMDA Type Subunit 2B). This TE was previously identified as “differentiated” among the isolates and is generally widespread in our six villages (Table 2). The other five intergenic elements are all Alus and are located on chr6:1257163, chr6:161283170, chr12:58367298, chr13:112866653 (eQTL in testis, skin and colon sigmoid) and chr18:26214257 (sQTL in testis, adipose visceral omentum, thyroid and breast mammary tissue).
3. Tobacco use (smoking): seven TEs were deemed significant (Figure 2, outer Manhattan plot), three of which are located in genes, namely the Alu on chr3:42856928, which acts as eQTL and sQTL in different tissues (including brain and lung) and is located in the gene *ACKR2* (Atypical Chemokine Receptor 2); the Alu on chr11:102654750 in *WTAPP1* (Wilms Tumor 1 Associated Protein Pseudogene 1), acting as eQTL in adrenal gland; and the Alu on chr12:129970510 in *TMEM132D* (Transmembrane Protein 132D). These last two Alus are mostly widespread in the six villages (Table 2) and were both identified as “differentiated” between the isolates (Table 2). The other four intergenic insertions are located on chr2:174296971 (Alu), chr2:188989149 (Alu, acting as sQTL in brain hippocampus and cerebellum), chr14:65796449 (Alu, acting as eQTL/sQTL in several tissues), and a LINE-1 on chr22:26454699.
4. A further association test with GEMMA was performed on the “smoking” condition by taking into account the number of cigarettes smoked per day and the number of years smoking. In total, six Alus were significantly associated with this phenotype, four of which were inserted in gene introns. The Alu on chr2:155232845, located in the gene *GALNT13* (Polypeptide N-Acetylgalactosaminyltransferase 13); the Alu on chr12:123580101, located in the gene *PITPNM2* (Phosphatidylinositol Transfer Protein Membrane Associated 2); the Alu on chr18:29519986, located in the gene *TRAPPC8* (Trafficking Protein Particle Complex Subunit 8); and the Alu on chr19:19350607, located in the gene *NCAN* (Neurocan).

**Figure 2.**
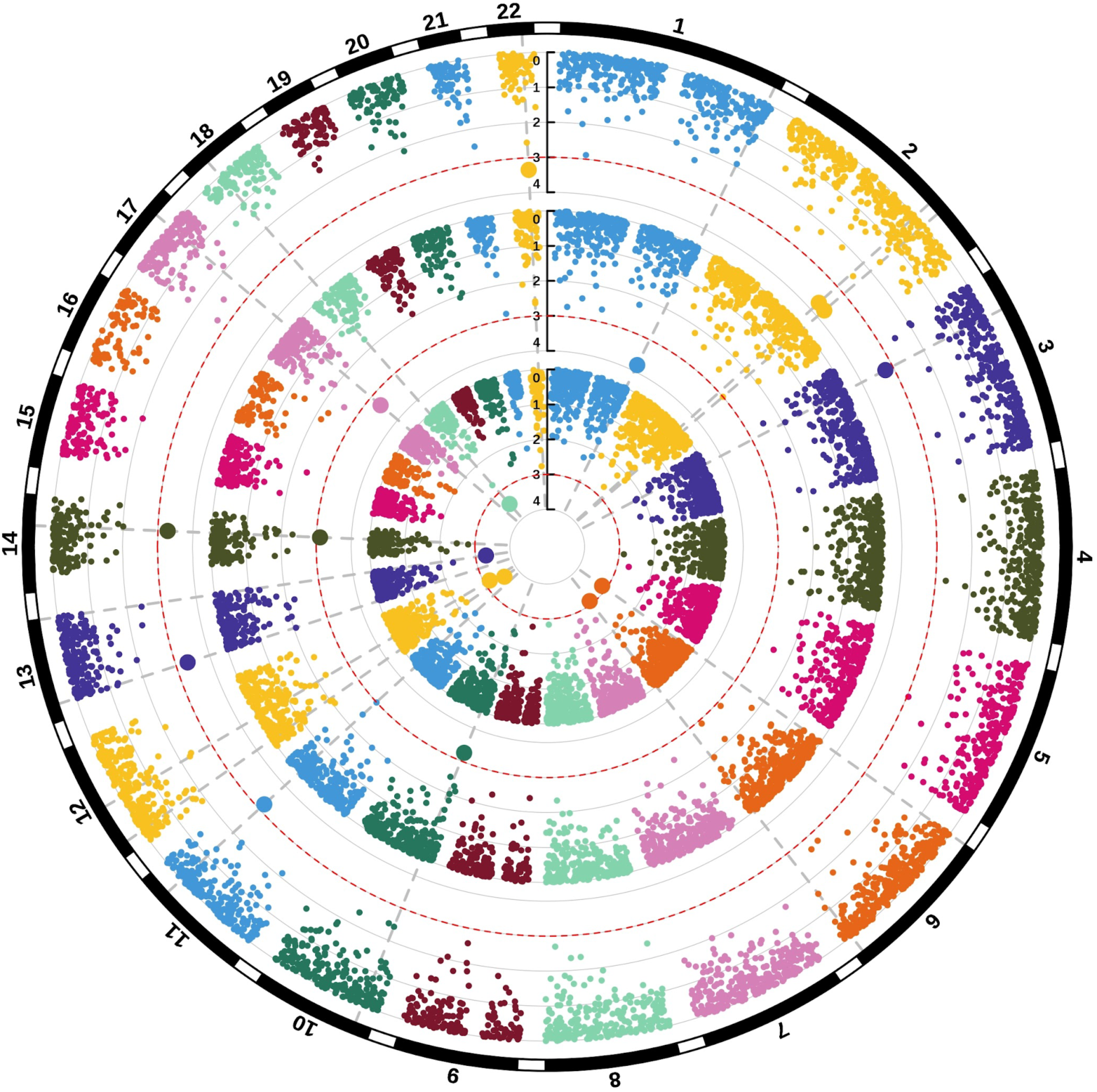
Circular Manhattan plots of the first three association tests (BMI, alcohol, smoking). The plot is read from the outside to the inside, in this order: smoking, BMI and alcohol. Greater dots over the red-dotted line (Wald’s p-value < 0.001) are the significant TEs for the association tests. Only autosomal chromosomes were represented as shown in the black circle outside the plots.

**Table 2.** Absolute genotype frequencies of the five Alus located in the genes *GRIN2B, WTAPP1, ACKR2, TMEM132D*, and *GALNT13*.

|  | Resia |  |  | Erto |  |  | Illegio |  |  | Sauris |  |  | San Martino |  |  | Clauzetto |  |  |
| --- | --- | --- | --- | --- | --- | --- | --- | --- | --- | --- | --- | --- | --- | --- | --- | --- | --- | --- |
|  | 0\0 | 0\1 | 1\1 | 0\0 | 0\1 | 1\1 | 0\0 | 0\1 | 1\1 | 0\0 | 0\1 | 1\1 | 0\0 | 0\1 | 1\1 | 0\0 | 0\1 | 1\1 |
| **GRIN2B** | 53 | 60 | 30 | 27 | 30 | 6 | 23 | 37 | 17 | 35 | 39 | 11 | 52 | 70 | 8 | 32 | 51 | 5 |
| **ACKR2** | 56 | 66 | 20 | 35 | 23 | 5 | 33 | 32 | 12 | 31 | 41 | 12 | 58 | 58 | 14 | 43 | 39 | 6 |
| **WTAPP1** | 104 | 37 | 1 | 59 | 4 | 0 | 65 | 11 | 1 | 76 | 9 | 0 | 119 | 11 | 0 | 77 | 11 | 0 |
| **TMEM132D** | 75 | 57 | 11 | 22 | 30 | 11 | 42 | 31 | 4 | 45 | 38 | 2 | 46 | 67 | 17 | 38 | 38 | 12 |
| **GALNT13** | 118 | 24 | 0 | 60 | 3 | 0 | 77 | 0 | 0 | 77 | 7 | 0 | 118 | 12 | 0 | 86 | 2 | 0 |

Among these genes, *TMEM132D*, and *GRIN2B* show evidence of genetic constraints using either pLI Score, RVIS, or SSC Score. Absolute genotype frequencies of these insertions are reported in Table 2. The Alus in the genes *SPAG9, PITPNM2, TRAPPC8* and *NCAN*, despite being significant, appear to be rare, therefore we did not report the genotype frequencies in Table 2 (the percentage of individuals who carry the insertion is 1.9% for SPAG9, 0.18% for *PITPNM2*, 0.26% for *TRAPPC8* and 0.19% for *NCAN*).

We finally reconstructed haplotypes around the above mentioned TEs and performed haplotype-based association tests as described in Methods. We obtained two significant results, both for the alcohol phenotype, namely the two intergenic Alus on chr6:1257163 and chr6:161283170. In both cases, the associated haplotype is characterized by the presence of the mobile element. The first haplotype included 19 SNPs and one Alu (p-value = 0.00164); the second is a haplotype with 7 SNPs and the TE (p-value = 0.000335).

## Discussion

The study of isolated communities is at the basis of population genetics research [47,48]. In fact, isolates yield genomes that show high homogeneity and are subject to similar environmental and cultural pressures, such as lifestyle habits, diet, sanitary conditions, and disease vectors. These populations are also an ideal subject to study the phenotypic effects of variants that were otherwise only marginally present in larger populations [48]. In this picture, Italian isolates are particularly important, mainly because of the peninsula’s central role in human migrations since prehistoric times and of the high number of genetically distinct isolated communities that have been established throughout history [49]. Polymorphic TEs, which have previously been used as both variability and susceptibility markers only in “general” populations [7,8,25], are here applied for the first time to human isolates. Using the Mobile Element Locator Tool [8] more than 12,000 polymorphic TEs were identified in the six villages of Friuli-Venezia Giulia. These TEs were used as genetic markers to obtain a first overview of their potential impact on diversity and disease susceptibility in isolated populations, in particular: 1) to study communities’ differentiation; 2) to explore the genetic variability of the isolates; 3) and to analyze their possible role as genetic variants underlying susceptibility to different behavioral traits or medical conditions (tobacco use, alcohol consumption, and BMI variations).

Firstly, after calculating allele and genotype frequencies of the identified TEs, we found that of 12,709 TEs, 3,987 (31.37%) have significantly different allele frequencies between the six isolates (Fisher’s exact test, p-value < 0.01), while the corresponding rate in European comparison populations is 53.78%. Considering the much lower geographic dimensions of FVG compared to Europe, these values suggest the presence of genomic structure among the isolates.

Then, TEs were used as markers for exploratory population analyses, such as PCA and Admixture, to look at the general diversity and ancestry of FVG isolates in the context of European genetic variability, as represented by the polymorphic TE content of European populations from 1KGP [8]. Our results show that FVG isolates tend to cluster amongst themselves (PC1 in Figure 1A, Figure 1C), compared to European populations; however some differentiation between the isolates is evident, particularly for Resia and some individuals from Clauzetto (PC2 in Figure 1B, Figure 1C), as well as Sauris and Illegio (PC3 in Figure 1B, Figure 1C). Instead, Erto, San Martino and most individuals from Clauzetto overlap with the other European populations. These results agree with previous SNP-based studies, according to which, Clauzetto is the least isolated village among the six FVG isolates [3]; at the same time Clauzetto, Erto and San Martino overlap to the considered European populations and have the lowest inbreeding coefficients among the villages [4]. The observed patterns of genetic variability and ancestry components could be explained by population structuring and genetic drift, a suggestion made also by previous works on the same dataset [3–5]. The observation of a strong correlation between SNP-driven results and TE-driven results in terms of population structure further highlights that the variability of polymorphic TE is mainly the result of demographic events.

To sum up, population structure analyses confirmed that on the whole, our populations show the typical marks of isolates also from the TEs point of view. As previously mentioned, due to their internal homogeneity both at genetic and social levels, isolates are ideal populations for performing genome-wide association studies. On the other hand, their relatively low census size implies a moderate number of available samples. More importantly, the presence of population structure is well known to induce false positives in association studies. However, the impact of the observed structure is probably moderate or at least not higher than in association studies at a country level, as suggested by exploratory population analyses (PCA, Admixture) and proportions of “differentiated” TEs. In addition, the usage of GEMMA should overcome distortions due to population structure, as confirmed by the fact that only a minority (3/22) of the associated variants show significant differentiation among isolates.

In this context, polymorphic TE insertions are particularly worthy of investigation, being potential risk variants for several medically relevant phenotypes, because of their innate ability to act as deregulators of gene networks [15]. Notably, the link between transposable elements and the health of the Central Nervous System is not new [26,27], with the effects of TEs being associated with stress, neurodegeneration, ageing, and drug abuse [25]. As such, TE markers can allow us to perform a first exploration of the medical susceptibility of individuals from the studied villages, by testing for association between TEs and phenotypes linked to behavioral and anthropometric traits.

Accordingly, we performed four association tests with GEMMA [35,36] in order to obtain a first overview of the polymorphic TEs that could underpin the variability of selected phenotypes, i.e. tobacco use, alcohol consumption, height and weight, from which we calculated body mass index (weight/height^2^). For tobacco use, two separate analyses were run, the first comparing smokers with non-smokers, the second only on smoker individuals, testing for the association between the number of cigarettes smoked per day and the number of years smoking. The GEMMA algorithm was selected due to its ability to take into account population structure, which is a typical feature of isolated populations. In addition, sex and age were introduced in the models as covariates: Manhattan plots are shown in Figure 2. Several TEs were deemed significant, some of which are located in known genes: an SVA (chr17:49150166) in the gene *SPAG9* (BMI variations); the Alu on chr3:42856928 in the gene *ACKR2*, the Alu on chr11:102654750 in *WTAPP1*, the Alu on chr12:129970510 in *TMEM132D* (tobacco use/smoking) and the Alu on chr12:14020945 in the gene *GRIN2B* (alcohol consumption). Interestingly, two of the results for the alcohol consumption phenotype were deemed significant also for the haplotype-based association test performed with Beagle [38]. In both cases, haplotypes including polymorphic TEs appear as significantly associated with the status “alcohol drinker”. As for the amount of cigarettes/number of years smoking, it resulted of interest the Alu insertion on chr2:155232845 in the gene *GALNT13*. Additionally, the insertions in *WTAPP1, TMEM132D*, and *GRIN2B* were also identified as “differentiated” when looking at genotype and allele frequencies between the isolates. Two of these genes (*TMEM132D* and *GRIN2B*) also show evidence of genetic constraints and thus should be prioritized in further investigations, as genes showing evidence of purifying selection in healthy individuals may be judged more likely to cause certain kinds of disease. For instance, the gene *GRIN2B* encodes a member of the ionotropic glutamate receptor superfamily and plays a major role in brain development and synaptic plasticity, with mutations in this gene often associated with neurodevelopmental disorders [50]. Moreover, variants of this gene have been associated with alcohol and tobacco consumption [51], general risk-taking behaviors [52], opioid dependence [53], and several neurological disorders such as schizophrenia [54] and Alzheimer’s disease [55]. Regarding tobacco use, the insertion in *ACKR2* (also known as D6) emerged as one of the most promising results, in fact the Alu acts as eQTL/sQTL in brain and lung tissues. The gene [56] controls chemokine levels and localization and is known to be involved in inflammatory responses [57]. Moreover, a work by Bazzan and colleagues [58] on chronic obstructive pulmonary disease (COPD) “demonstrates an increased expression of the atypical chemokine receptor D6 in peripheral lung from smokers with COPD but not in smoking subjects who did not develop the disease and nonsmoker control subjects”. Furthermore, *TMEM132D*, encoding for a transmembrane protein, has already been associated with many neurological disorders such as anxiety and panic disorders [59] and general behavioral disinhibition, including alcohol consumption and dependence, illicit drug use, and nicotine use [60]. Lastly, an Alu inserted in the gene *GALNT13* was found to be highly associated with the prolonged use of tobacco (number of cigarettes smoked daily/number of years of smoking). Mutations in *GALNT13*, which normally encodes for Polypeptide N-Acetylgalactosaminyltransferase 13, a transferase linked with the metabolism of proteins and the glycosylation of mucins [61] also expressed in the Purkinje cells of the developing brain [62], have been associated with nicotine use [51], severe comorbidity between nicotine dependence and major depression [63]. Furthermore, *GALNT13* expression has been found to be increased in lung cancers [64], suggesting yet another deep link between the identified gene insertions and genes involved in brain development, possible neurological/physical diseases and addiction-seeking behaviors.

Polymorphic transposable elements emerge as a compelling avenue for elucidating human genetic diversity. The innovative utilization of polymorphic TEs as markers for genetic variability within isolated communities represents an unprecedented methodological advancement. This study demonstrates the utility of polymorphic TEs in effectively encapsulating genetic variability and historical contexts among isolates, substantiated by congruent outcomes with prior investigations relying on single nucleotide variants [3–5]. While progress has been made, the comprehensive impact of transposable elements on the human genome remains incompletely understood, as does the cascade of effects on diverse phenotypes. This investigation identifies numerous TE insertions correlated with specific phenotypes, such as substance use and metabolic disorders. It is imperative to underscore the exploratory nature of our analyses, necessitating further empirical validation to establish definitive causal links between these insertions and medical susceptibility. Nevertheless, the identified insertions stand as pivotal points of interest, providing a foundational platform for subsequent research. In the context of isolated communities, these populations serve as invaluable "laboratories", affording unique insights into the influence of transposable elements on physical, psychological, and behavioral traits. Consequently, prospective studies should prioritize the validation of identified variants and engage in selection analyses to discern potential instances of natural selection within these isolated populations. This forward-looking research agenda holds significant promise for advancing our understanding of the intricate interplay between transposable elements and human phenotypic traits.

## Materials and Methods

The dataset used in this study was generated in 2008 [3–5] from the sampling of 611 individuals from six geographically and historically isolated villages in the Friuli-Venezia Giulia region of North-Eastern Italy, namely Sauris, Illegio, Resia, Erto, Clauzetto and San Martino del Carso (Figure 3). Since a few of the individuals present in the dataset were duplicates (specifically, 22 individuals from Resia), and three missed village information, they were removed, leading the total number of analyzed individuals to 586. During the sampling, subjects were asked to fill out an anamnesis form to acquire more data on their general health and lifestyle habits. Phenotypic data on more than 70 phenotypes was collected, also including food preferences, olfactory perception, gustatory perception and anthropometric measures. Since the form was administered at individual discretion, missing rates vary wildly between phenotypes and individuals. We chose to focus solely on phenotypes exhibiting a missing rate of less than 10% in our dataset, thus the trait included in our analysis were sex, age, alcohol consumption, smoking, as well as height and weight, from which we calculated the corresponding BMI (weight/height^2^). Phenotypes linked to specific diseases or health conditions, such as the occurrence of diabetes, displayed a missing rate exceeding 40%, and as such we chose to not include them in our analyses.

**Figure 3.**
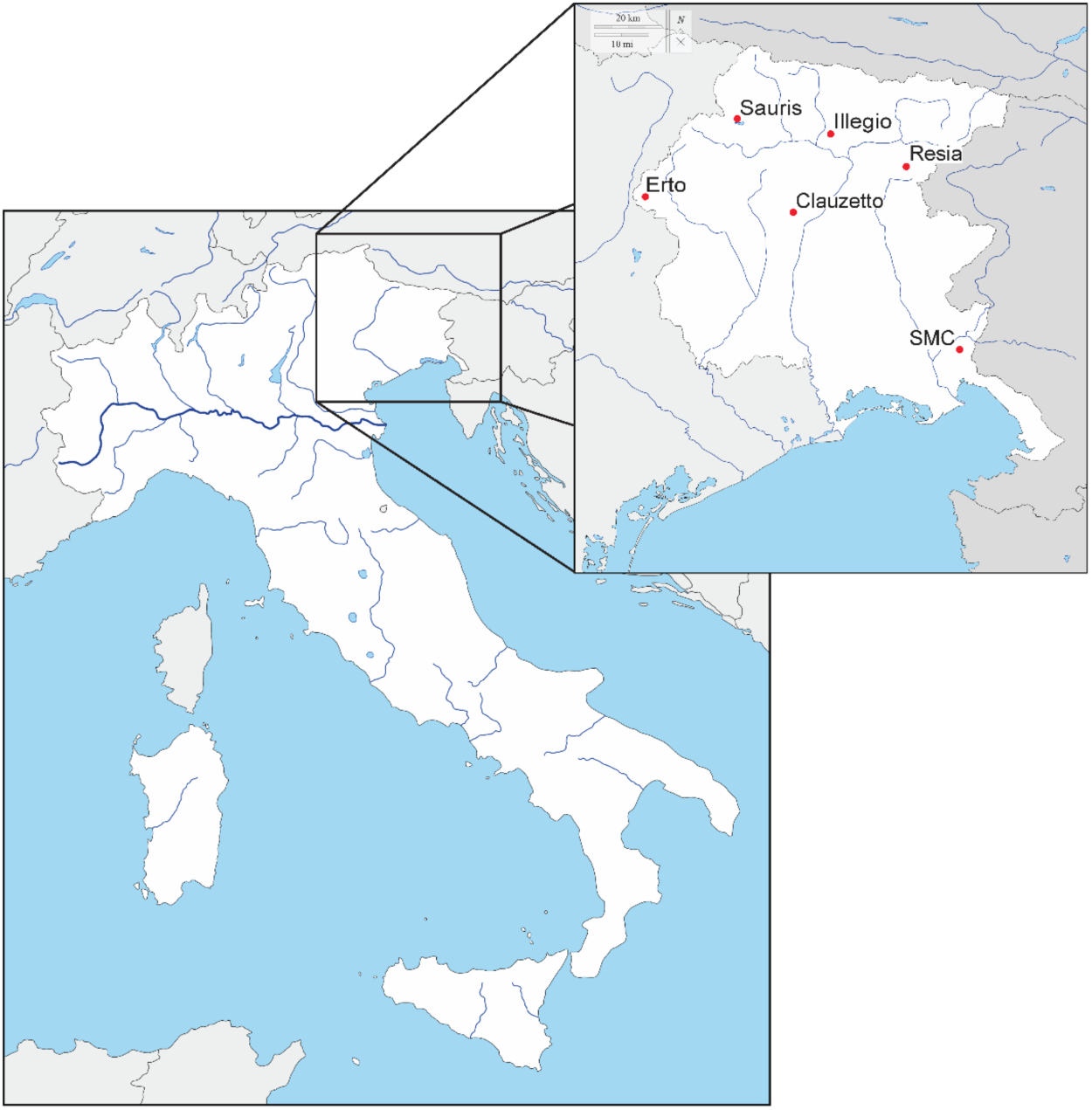
Location of the six isolates in Friuli-Venezia Giulia, north-east of Italy (SMC = San Martino del Carso).

Genomes were scanned in search of non-reference polymorphic TEs (Alus, LINE1s, and SVAs), using the Mobile Element Locator Tool (MELT) v2.2.2 [8]. The WGS data was aligned with *bwa* to the human reference HumanG1Kv38, and the aligned reads were used as input for the Mobile Element Locator Tool (MELT). For the calling process, we used the MELT mobile element reference sequences and the collection of insertion sites discovered in Phase III of the 1000 Genomes Project (1KGP) as analysis priors. After the identification of these TEs, a self-customized Python script was applied to the resulting *vcf* files to calculate both allele and genotype frequencies of each TE for all the isolated villages. Allele frequencies were then analyzed for significant differences between villages with Fisher’s exact test, using a significant threshold of nominal p-value < 0.01 (“differentiated” TEs).

MELT provides gene names in RefSeq format: therefore, RefSeq accession numbers were converted to their respective Official Gene Symbol using the Database for Annotation, Visualization and Integrated Discovery (DAVID) (https://david.ncifcrf.gov/) [30], taking into consideration the specific gene region TEs were inserted in (Intron, Exon, Promoter, Terminator, 5’ UTR and 3’ UTR).

To compare TE diversity of the isolates with other human populations, we built a new dataset consisting of polymorphic TEs identified with MELT [8] that were present both in the six isolates and in the populations of the 1000 Genomes Project. This newly merged dataset contained a total of 2,814 genetic loci for 3,090 individuals from 32 populations. The populations were divided into 6 groups based on geographic macro areas, consistent with the super populations of 1KGP [31], i.e. Africa (AFR), America (AMR), East Asia (EAS), Europe (EUR), South Asia (SAS), plus the isolates from Friuli-Venezia Giulia (FVG).

TEs were coded as single nucleotide variants, substituting the insertion with a nucleotide base that was non-complementary to that of the other allele in the genotype file. Information about the true nature of each insertion was kept in the original *vcf* file. Variants were then filtered with PLINK v1.9 [32] as follows: 1) Removal of insertions located on sexual chromosomes or mitochondrial genome insertions, to retain only autosomal variability and removal of duplicates, using the --*exclude* option. 2) Exclusion of individuals and variants with > 1% missing data with the commands --geno 0.01 (for variants) and --mind 0.01 (for individuals). 3) Removal of variants that did not respect the Hardy-Weinberg Equilibrium (HWE) with the option --hwe, setting a significant threshold of 0.01 using a Bonferroni Correction for multiple testing (threshold= 0.01/number of variants). 4) Removal of variants with a minor allele frequency < 0.01 (--maf 0.01). 5) Removal of closely related individuals with an Identity by Descent (IBD) estimate higher than 0.25, using the --genome option to calculate the pairwise IBD estimates between every couple of individuals and --remove to exclude one of the two related individuals. Therefore, the final filtered dataset was made of 1,703 variants shared among 3,087 individuals.

The generated dataset was then used to perform a series of analyses on TE insertions from the six isolates when compared to 1KGP groups. Both a Principal Component Analysis (PCA) and Admixture analysis were applied: PCA was performed after the conversion from the PLINK format (*bed, bim, fam*) with the *convertf* and *smartpca* tools of the EIGENSTRAT v6.0.1 package [33]. Admixture was implemented with the ADMIXTURE tool [34], testing between 2 and 23 potential ancestry components (K) and performing 50 iterations of each run to minimize the estimation error and maximize the *log-likelihood* of each ancestry estimate.

We then compared FVG isolates with other European populations, subsampling the original 1KGP dataset as follows: Utah residents with North-Western European ancestry (CEU), Finnish in Finland (FIN), British in England and Scotland (GBR), Iberian populations in Spain (IBS), Tuscans in Italy (TSI). PCA and Admixture analyses were implemented using the above approach, the only difference being that we tested a number K of putative ancestry components between 2 and 12.

As introduced in the “background” section, individuals were asked to fill out an anamnesis form, including information on their health status and lifestyle habits. Three phenotypes were selected to perform association studies between polymorphic TEs and the considered traits: tobacco use, alcohol consumption, and body mass index (the latter was calculated as weight/height^2^). The association studies were performed with the software GEMMA [35,36], using a genome-wide (GWAS) like approach. In particular, we applied for all the considered phenotypes a univariate linear mixed model (uvLMM) for association tests between a marker, a chosen phenotype, and any chosen covariates, while also correcting for the potential presence of population stratification (indeed a typical feature of isolates), and estimating genetic correlation among phenotypes [36]. GEMMA was applied to the full FVG dataset (12,709 TEs and 586 individuals) and three separate uvLMM association analyses were performed, using sex and age as covariates: 1) BMI; 2) a binary alcohol drinker/non-drinker variable (set as “1” for drinker individuals and “0” for non-drinkers); 3) a binary smoker/non-smoker variable (using “1” for smokers and “0” for non-smokers). A fourth association test on the smoker individuals was performed to evaluate the possible association between polymorphic TEs and the number of cigarettes smoked per day/number of years smoking. Variables were tested using Wald’s test with a significant threshold of p-value = 0.001; Manhattan plots with significant results have been obtained with the *CMplot* R package (https://github.com/YinLiLin/CMplot) [37]. To better assess the importance of the associated TEs, we also performed a haplotype reconstruction/association test procedure on the significant variants from the alcohol and smoking tests detected with GEMMA using Beagle [38] (this software performs association tests only on binary traits). First, we selected regions of interest (10kb upstream and downstream the significant TE, for a total of 20kb) with VCFtools [39] and phased those regions with the software Beagle v5.1. The obtained *vcf* files were converted into the Beagle format with vcf2beagle (https://faculty.washington.edu/browning/beagle_utilities/) and the case status “smoking” or “alcohol” was included in the second row of the *bgl* files. Lastly, the association test on the reconstructed haplotypes was performed with Beagle v3.3.2 and the significant results were checked with the cluster2haps utility. In order to investigate a possible function for the identified TEs, we then cross-checked the significant results with the lists of polymorphic TEs acting as expression/alternative splicing quantitative trait loci produced by Cao and colleagues [40]. For each gene analyzed we collected measures of genetic constraints such as pLI (probability of loss of function intolerance) [41], RVIS (Genic Intolerance) [42] and SSC score (Singletons Score) [43] for prioritization. We considered as constrained those genes with pLI > 0.9 or RVIS < -0.43 or SSC score < -2.

## Data Availability

Genetic Data of isolated populations are available in the European Genome-phenome Archive (EGA).

https://www.ebi.ac.uk/ega/studies/EGAS00001000252

https://www.ebi.ac.uk/ega/studies/EGAS00001001597

https://www.ebi.ac.uk/ega/datasets/EGAD00001002729

## Data access

Genetic Data of isolated populations are available in the European Genome-phenome Archive (EGA) at the following links. BAM files: https://www.ebi.ac.uk/ega/studies/EGAS00001000252 (accessed on 21st Februrary 2024); sample list, vcf files: https://www.ebi.ac.uk/ega/studies/EGAS00001001597 (accessed on 21st Februrary 2024); https://www.ebi.ac.uk/ega/datasets/EGAD00001002729 (accessed on 21st Februrary 2024).

## Human Ethics and Consent to Participate

Not applicable.

## Authors contributions

A.B. and M.M. conceived the study. G.M. (Giorgia Modenini), G.M. (Giacomo Mercuri) and P.A. performed all the *in silico* analyses and plotted results. G.G.N., A.S., P.T., B.S., A.P., G.P., M.P.C., G.G. and P.G. collected data and published them on the European Genome-phenome Archive. G.M. (Giorgia Modenini), G.M. (Giacomo Mercuri), P.A., A.B. and M.M. wrote the manuscript. All Authors read and approved the manuscript.

